# The Austrian MS Database and the Austrian MS Cohort – a national effort towards data harmonization and prospective data collection

**DOI:** 10.1101/2025.02.17.25322422

**Authors:** Gabriel Bsteh, Fabian Föttinger, Markus Ponleitner, Klaus Berek, Franziska Di Pauli, Bettina Heschl, Sebastian Wurth, Florian Deisenhammer, Christian Enzinger, Thomas Berger, Michael Khalil, Harald Hegen, the Austrian Multiple Sclerosis Database and Austrian MS cohort study group

## Abstract

**Background:** A variety of disease-modifying therapies (DMTs) are now available for multiple sclerosis (MS), each with distinct efficacy and risk profiles. However, the clinical course of MS varies significantly both within and between individuals, and the optimal treatment strategy remains uncertain at the group and individual levels. Addressing questions about treatment strategies through traditional randomized controlled trials is unrealistic due to the large sample sizes and high costs required. Instead, large-scale multicenter registries and well-characterized prospective observational cohorts offer a feasible approach to generating meaningful insights. For valid results, such registries and cohorts require harmonization across centers to support comprehensive, standardized, user-friendly data collection while meeting data protection standards and ensuring quality control.

**Objective:** This project aims to establish standardized, nationwide MS data collection in Austria.

**Methods:** The project consists of five key components: i) harmonization of data collection, ii) creation of infrastructure for data sharing, iii) retrospective harmonized data collection (Austrian MS Database, AMSD), iv) prospective harmonized data collection (Austrian MS Cohort, AMSC), and v) aggregated analyses.

**Results:** A comprehensive set of harmonized common data elements (CDE) comprising clinical and paraclinical data was developed and a common data collection infrastructure was generated using the web-based Research, Documentation, and Analysis platform (webRDA), an innovative data capture, processing, and analysis system provided by the Medical University of Vienna offering pseudonymized storage of data supported by a robust permissions system sufficing legal data protection and ethical requirements.

The AMSC is set up as a standardized prospective collection of demographic, clinical, epidemiological, psycho-socio-economic, MRI, and OCT data as well as body fluids.

**Conclusion:** The AMSD and AMSC will facilitate the evidence-based development of prognostic biomarkers, individualized therapy strategies and treatment sequences based on a high-quality, population-based dataset of more than 8,000 people with MS.

## Background

Multiple Sclerosis (MS) is an immune-mediated, chronic inflammatory, demyelinating disease of the central nervous system and is the leading cause of neurological disability in young adults [1]. Over the past three decades, an ever-growing armamentarium of disease-modifying therapies (DMTs) has emerged, enabling effective suppression of disease-activity [2,3]

These DMTs vary considerably in terms of efficacy and associated risks. With the availability of high-efficacy DMTs, the goal of MS therapy has shifted from simply managing disease course (reducing relapses, disability progression, and MRI disease activity) to achieving suppression of disease activity below detectable levels [4].

However, it is essential to recognize that MS exhibits extremely variable intra- and inter-individual courses. Some people with MS (pwMS) experience highly active disease with breakthrough activity even under high-efficacy DMTs (HE-DMT), while others have a mild disease course that may not necessitate a potentially risky, psychologically burdensome, and costly DMT [5]. Conversely, a growing perspective advocates for HE-DMT in the majority, or even all, pwMS. The increasing array of therapeutic options also raises questions about the optimal sequencing of these therapies.

Currently, there is a lack of direct prospective comparative studies between individual therapies, therapy sequences, and treatment strategies to address these questions in an evidence-based manner. A significant challenge is that the clinical trials for DMTs, conducted under strict inclusion criteria, exclude a relevant proportion of pwMS [6]. Furthermore, the impact of DMTs on various aspects of MS pathology and the influence of factors such as lifestyle, psycho-socio-economic background, comorbidities, and aging are not addressed in these studies.

Due to the enormous sample sizes and associated costs, conducting such studies with the classical randomized controlled trial design is impractical. Thus, the only feasible approaches for generating meaningful insights in this area lie in large-scale, multicenter registries and well-characterized prospective long-term observational cohorts combining the systematic acquisition of clinical data, imaging and laboratory measures.

However, for such projects to yield valid results, two essential prerequisites must be met: a harmonized, universally accepted approach for data collection, wherein all contributors collect and document data in a consistent manner, and a data infrastructure that allows standardized, comprehensive, and user-friendly data collection while adhering to data protection and security requirements and enabling quality control. Finally, such registries and databases must be designed to accommodate contributions from as many eligible centers as possible to maximize sample size and population diversity, and the collected data should be accessible for analysis by a broad research community (data sharing). In the field of MS, there have been several such projects such as various national MS registries and the well-known "MS Base" registry [7]. However, these projects cover a highly limited set of common variables (basic demographic data, clinical course with relapses and EDSS, as well as type, timing, and duration of DMTs, only sometimes including even basic paraclinical data such as the number and dynamics of MRI lesions). Furthermore, these datasets were not harmonized, meaning they were collected and documented in different ways and are subject to only rudimentary quality control, resulting in compromised data quality and limited data sharing capability.

In Austria, the nationwide network of certified MS centers, established by the Austrian Society of Neurology (ÖGN), provides high-quality care and documentation. However, the currently established Austrian MS Therapy Registry (AMSTR) only includes pwMS receiving a DMT (excluding interferon beta or glatiramer acetate formulations), leaving a significant portion of the more than 14,000 pwMS in Austria unrecorded [8,9].

In 2016, the Medical Universities of Vienna, Innsbruck and Graz initiated a multi-center biomarker project in MS with four specific aims: i) to establish and maintain a long-term cohort of pwMS in Austria, ii) to systematically follow these pwMS with standardized collection of demographic, clinical, MRI and body fluid data, and iii) to uphold and enhance the high standard of MS care in Austria by evaluating the long-term efficacy and safety profiles of available DMT.

Based on these previous efforts and collaborative structures, this research project aims i) to establish a harmonized, nationwide collection of a broad array of real-world data on pwMS in Austria (Austrian MS Database, AMSD) and ii) to evolve the prospective long-term observational comprehensive cohort study of demographic, clinical, epidemiological, psychosocio-economic, MRI, and OCT data as well as body fluids (Austrian MS Cohort, AMSC), in order to facilitate nested projects on prognostic indicators, biomarkers, individualized therapy strategies and treatment sequences.

## Methods

The project consists of five key components:

1. Harmonization of data collection
2. Infrastructure creation for collection, management and sharing of data
3. Retrospective and prospective harmonized collection of real-world data (AMSD)
4. Prospective harmonized collection of comprehensive high-quality data (AMSC)
5. Analyses of aggregated data

### Harmonization of data collection

The harmonization of data collection was coordinated by the AMSD core centers comprised of the Departments of Neurology of the Medical Universities of Vienna, Innsbruck and Graz. The core centers developed a draft of common data elements (CDE), which were then discussed with representatives from MS clinics of all participating sites and refined over multiple iterations into a final set of CDEs including precise definitions of documentation and coding of data. This final dataset serves as the harmonized foundation for data collection within the AMSD and encompasses patient data (demographics, family history, education, occupation, social environment, consumption of alcohol and nicotine), clinical data (MS disease course, initial symptoms, relapses, relapse remission, expanded disability status scale [EDSS], comorbidities, concomitant medications, pregnancies, vaccination status), paraclinical data (cerebral/spinal magnetic resonance imaging [MRI], optical coherence tomography [OCT], serum and cerebrospinal fluid parameters/biomarkers) and treatment (relapse treatment, disease-modifying therapies [DMT] including side effects, symptomatic therapies) (see Figure 1). A complete summary of AMSD-CDE is provided in the supplements (Supplemental File 1).

**Figure 1.**
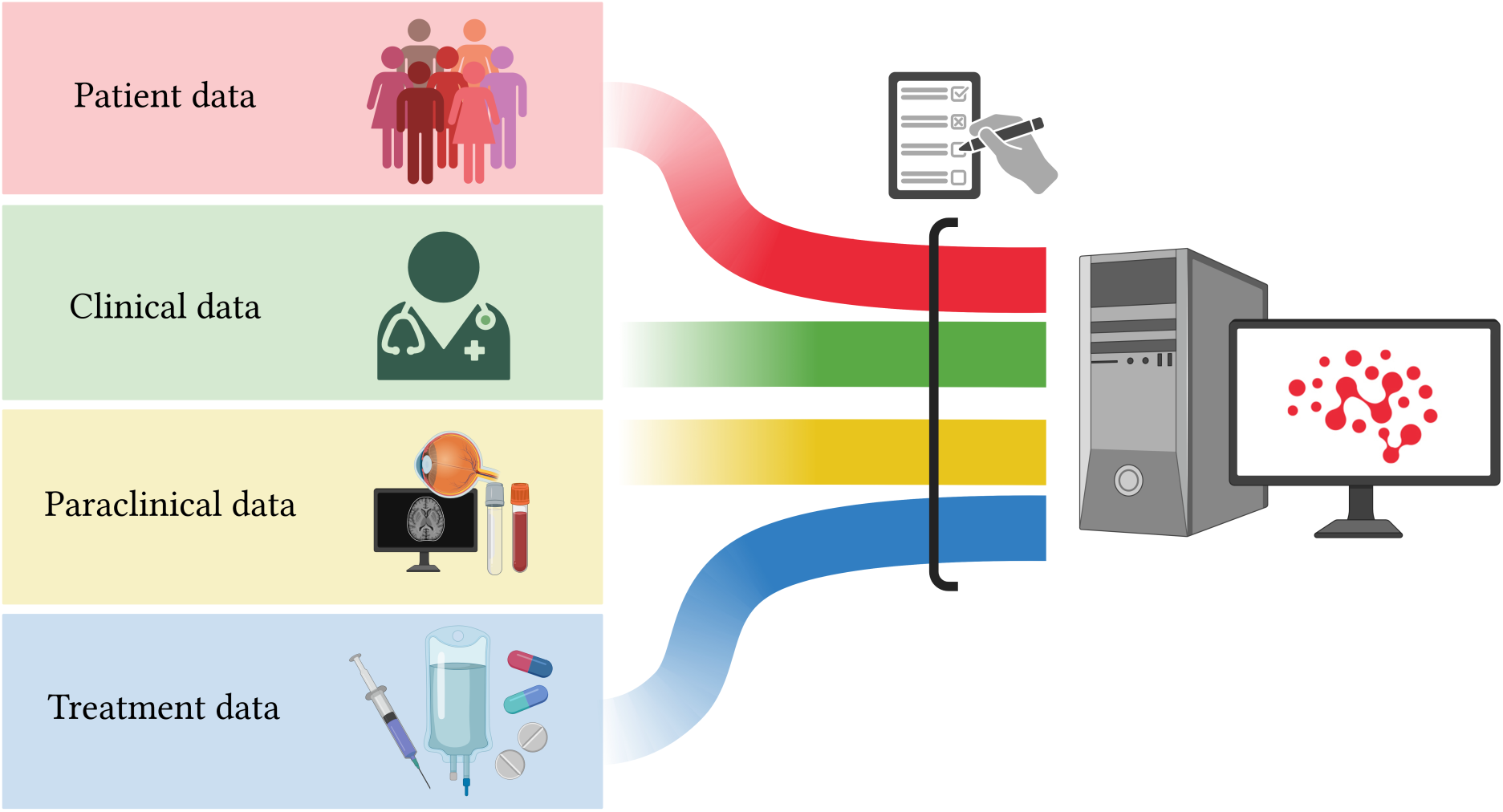
AMSD Overview.

### Infrastructure for data sharing

The data infrastructure is developed as a web-based Research, Documentation and Analysis (RDA) database, a non-commercial platform supported by Medical University of Vienna enabling collection of research-relevant data in specifically designed modular registries [9]. This data pool is continuously and automatically supplied with specifically selected documented routine data, which are fed into RDA registries. The combined use of both data sources ensures enhanced scientific utilization and includes functionalities such as form-based documentation, data collections for multicenter registries, patient-specific and pseudonymized documentation, and the automated transfer of data from various sources. From a data protection perspective, it is particularly noteworthy that the RDA offers pseudonymized web-based documentation for multicenter registries, supported by a robust permissions system (including read, edit, and query rights, along with the logging of all access events).

The AMSD-RDA comprises all agreed AMSD-CDE, which creates a virtual database integrating data from all participating centers into a unified data model.

Each core center has – as well as each future participating center will have – reading and writing access only to the patient data entered by its own center. Centers not able to use the web-based RDA due to local legal or technical restrictions can also use an off-line version of the AMSD using the same CDE to collect data. Each participating MS center signs a data-sharing agreement that ensures data protection and grants end users reading and writing access exclusively to the patient data entered by their respective center.

### Retrospective harmonized data collection and data management (AMSD)

At all participating MS centers, the AMSD is populated retrospectively using pre-existing local databases and patient records, as well as prospectively following inclusion into the AMSD. The dataset includes all patients who meet the following criteria:

1. Diagnosis of multiple sclerosis according to the McDonald criteria, in the version applicable at the time of initial diagnosis (2001/2005/2010/2017) [10–13].
2. Initial diagnosis on or after January 1, 2001.

Data collection is conducted in a pseudonymized form, with each patient assigned a unique, center-specific sequential ID number. The datasets can only be linked to identifiable patient information through a subject log, which remains exclusively at each center.

Retrospective data collection is performed by trained personnel using the existing database from the core centers and who are supervised by the AMSD project core team to ensure standardized and high-quality data collection [14]. Quality assurance is provided through double data extraction on a 5% sample of entries and a plausibility check by the AMSD project core team on another 5% sample.

The AMSD was approved by the ethics committee of the coordinating center at the Medical University of Vienna (ethical approval number: 1668/2023) as well as the ethics committee of the Medical University of Innsbruck (approval number: 1050/2023) and Graz (approval number: 31-432 ex 18/19). As data are extracted from pre-existing local databases and patient records obtained in routine practice, the need for written informed consent from study participants was waived by the ethics committee.

### Prospective harmonized data collection (Austrian MS Cohort, AMSC)

The Austrian MS Cohort (AMSC) is an evolution of the previous research projects initiated between the core centers in Vienna, Innsbruck and Graz, which began recruitment in 2016, with four specific aims: i) to establish and maintain a long-term cohort of pwMS in Austria, ii) to systematically follow these pwMS with standardized collection of demographic, clinical, epidemiological, psycho-socio-economic, MRI, and OCT data as well as body fluids, and iii) to uphold and enhance the high standard of MS patient care in Austria by evaluating the long-term efficacy and safety profiles of available DMT for MS.

To be included in the AMSC, individuals must be diagnosed with either relapsing-remitting MS (RRMS), secondary-progressive MS (SPMS), or primary-progressive MS (PPMS) according to the 2017 revised McDonald criteria [13, 15]. Patients diagnosed with clinically isolated syndrome (CIS), radiologically isolated syndrome (RIS), neuromyelitis optica (NMOSD) and myelin oligodendrocyte glycoprotein antibody-associated disease (MOGAD) may also be included [16–18].

Patients are enrolled in the AMSC only after signing written informed consent. Every effort is made to minimize dropouts. If a patient relocates within Austria, the original center coordinates a consultation with the nearest participating center.

Data collected within the AMSC is entered and managed within the RDA framework. Data quality is subject to several automatic and manual internal quality checks including annual quality control checks of a random sample of 5% records and another random sample of 10% newly added records. When inconsistencies are observed, queries are sent to the respective center until the discrepancy is successfully solved.

The AMSC was approved by the ethics committee of the coordinating center at the Medical University of Vienna (approval numbers: 2195/2016 and 1668/2023), as well as the ethics committee of the Medical University of Innsbruck (approval number: 1050/2023) and Graz (approval number: 31-432 ex 18/19) and will be approved as well by independent ethics committees at each participating center.

#### Collection of demographic, epidemiological, clinical and psycho-socio-economic data

At baseline, variables collected include sex, date of birth, height, weight, ethnicity, family history, pregnancy history, consumption of alcohol and nicotine, date and type of first MS symptom, date and number of relapses, type of relapses, relapse treatments and remission, date of diagnosis, MS disease course, date of progression onset (if applicable), current and prior DMT, as well as concomitant medical conditions, medications and vaccination status. Standardized clinical assessments, including EDSS calculation with functional system scores, Timed 25-Foot Walk Test (T25FW), 9 Hole Peg Test (9HPT) and Symbol Digit Modalities Test (SDMT) are conducted by certified raters [19–21].

In addition, while not mandatory within the AMSC, we aim to obtain the following clinical scales and questionnaires at baseline: Beck Depression Inventory (BDI) and/or the Hospital Anxiety and Depression Scale (HADS) to quantify depression and/or anxiety, Fatigue Scale for Motor and Cognitive Functions (FSMC) to quantify fatigue, Multiple Sclerosis Impact Scale (MSIS-29) to quantify MS-related quality of life, the PeRiCoMS test battery to quantify personality, risk awareness, and coping strategies, and the AMSC socioeconomic inventory (SCOPE-MS AT) to retrieve data on socioeconomic and occupational status, housing and living conditions, healthcare access and medical expenses, support network and social life, health literacy and self-management, psychosocial and emotional well-being [22–26].

Each patient is followed up every 6 or 12 months ±45 days, as determined by the treating physician. At each visit, data on the occurrence of relapses, disability worsening (as measured by EDSS, T25FW, 9HPT, SDMT), initiation or interruption of DMT, DMT-related adverse events, weight, additional medical conditions, and concomitant medications are recorded. In addition, while again not mandatory within the AMSC, we aim to obtain BDI and/or HADS, FSMC, MSIS-29 at each follow-up visit and within 3-6 months after each DMT start (rebaseline). The PeRiCoMS and SCOPE-MS AT are aimed to be performed biennially and at rebaseline.

#### Magnetic resonance imaging

MRI acquisition is not mandatory within the AMSC. However, our goal is to obtain cranial MRI annually to biennially from as many patients as possible (ideally all). In addition, we aim to conduct cranial MRI within 3 to 6 months after DMT initiation or change (rebaseline). All scans should be performed within ±28 days of clinical data and sample collection [27].

A state-of-the-art MRI protocol, established at the core centers Vienna, Innsbruck and Graz already in 2016 and to be agreed upon by all contributing AMSC centers, includes high-resolution isotropic T1 (3D) without gadolinium (Gd) and post-intravenous Gd contrast, 3D-FLAIR isotropic (or 2D if 3D is not feasible), and susceptibility-weighted imaging (SWI). We aim to perform MRI scans primarily at 3T magnet strengths, but 1.5T are accepted as well. Advanced MRI sequences (e.g., DTI, MTR, etc.) may be included as part of nested projects. MRI readings are performed and stored locally at the AMSC core centers and can be uploaded securely for quality control, centralized storage, backup, and standardized analyses within the Neurodesk platform [28].

#### Optical Coherence Tomography

OCT is not mandatory within the AMSC. However, our goal is to perform spectral-domain OCT in accordance with the Austrian Network for OCT in MS (AN-OCT-MS) consensus [29]. for each AMSC participant at diagnosis (if applicable), before each DMT start, within 3-6 months after each DMT start (rebaseline) and then annually to biennially. Routine parameters obtained include peripapillary retinal nerve fiber layer (pRNFL) and ganglion cell and macular inner plexiform layer (GCIPL) thicknesses separate for each eye with documentation of optic neuritis history. All examinations are checked for sufficient quality using OSCAR-IB criteria. Scans from patients/eyes with diagnoses of ophthalmologic (e.g., myopia greater than −6 diopters, optic disc drusen, glaucoma), neurologic, or drug-related causes of retinal damage not attributable to MS are excluded [29, 30]. OCT readings are performed and stored locally at the AMSC core centers and can be uploaded securely for quality control, centralized storage, backup, and standardized analyses by the Vienna Reading Center (VRC) [31].

#### Biobanking

At each AMSC visit, serum and plasma samples are collected via venipuncture for biobanking according to standardized guidelines. For the purpose of genotyping, DNA-EDTA whole blood samples are collected at least once from each AMSC participant. During lumbar punctures performed as part of routine clinical care, cerebrospinal fluid (CSF) samples are also collected for biobanking. Serum, plasma, DNA-EDTA whole blood, and CSF samples are then immediately stored at −80°C in biobanks established at the core centers (Vienna: approval number 2195/2016, Innsbruck: approval number 1050/2023, Graz: approval number 31-432 ex 18/19) in accordance with international consensus guidelines [32, 33]. Control groups will be defined based on specific study questions but following standardized guidelines [34].

**Table 1.** Austrian Multiple Sclerosis Cohort Overview.

|  | Screening | Baseline | Follow-up<br>every 6-12<br>months | Follow-up<br>every 12-24<br>months | Rebaseline<br>(3-6 months<br>after<br>DMT<br>start/change) |
| --- | --- | --- | --- | --- | --- |
| Written Informed Consent | X |  |  |  |  |
| Inclusion criteria | X |  |  |  |  |
| History |  | X |  |  |  |
| Ethnicity |  | X |  |  |  |
| Family history |  | X |  |  |  |
| MS disease course |  | X | X | X | X |
| Relapses |  | X | X | X | X |
| DMT |  | X | X | X | X |
| Concomitant Medication |  | X | X | X | X |
| Concomitant Conditions |  | X | X | X | X |
| Vaccination status |  | X | X | X | X |
| Pregnancy history |  | X | X | X | X |
| Height |  | X | X | X | X |
| Weight |  | X | X | X | X |
| EDSS |  | X | X | X | X |
| SDMT |  | X | X | X | X |
| T25FW |  | X | X | X | X |
| 9HPT |  | X | X | X | X |
| BDI or HADS (optional) |  | X | X |  | X |
| FSMC (optional) |  | X | X |  | X |
| MSIS-29 (optional) |  | X | X |  | X |
| PeRiCoMS (optional) |  | X |  | X | X |
| AMSC socioeconomic<br>inventory (SCOPE-MS-AT, optional) |  | X |  | X | X |
| MRI |  | X |  | X | X |
| OCT |  | X |  | X |  |
| Biobank |  | X | X | X | X |

AMSC: Austrian Multiple Sclerosis Cohort. BDI: Beck Depression Inventory. DMT: disease-modifying treatment. EDSS: expanded disability status scale. FSMC: Fatigue Scale for Motor and Cognitive Fatigue. HADS: Hospital Depression and Anxiety Scale. MRI: magnetic resonance imaging. MSIS-29: Multiple Sclerosis Impact Scale. OCT: optical coherence tomography. PeRiCoMS: Personality, risk perception and coping in MS battery. SDMT: symbol digit modalities test. T25FW: Timed 25-foot walk test. 9HPT: 9-hole peg test.

**Figure 2.**
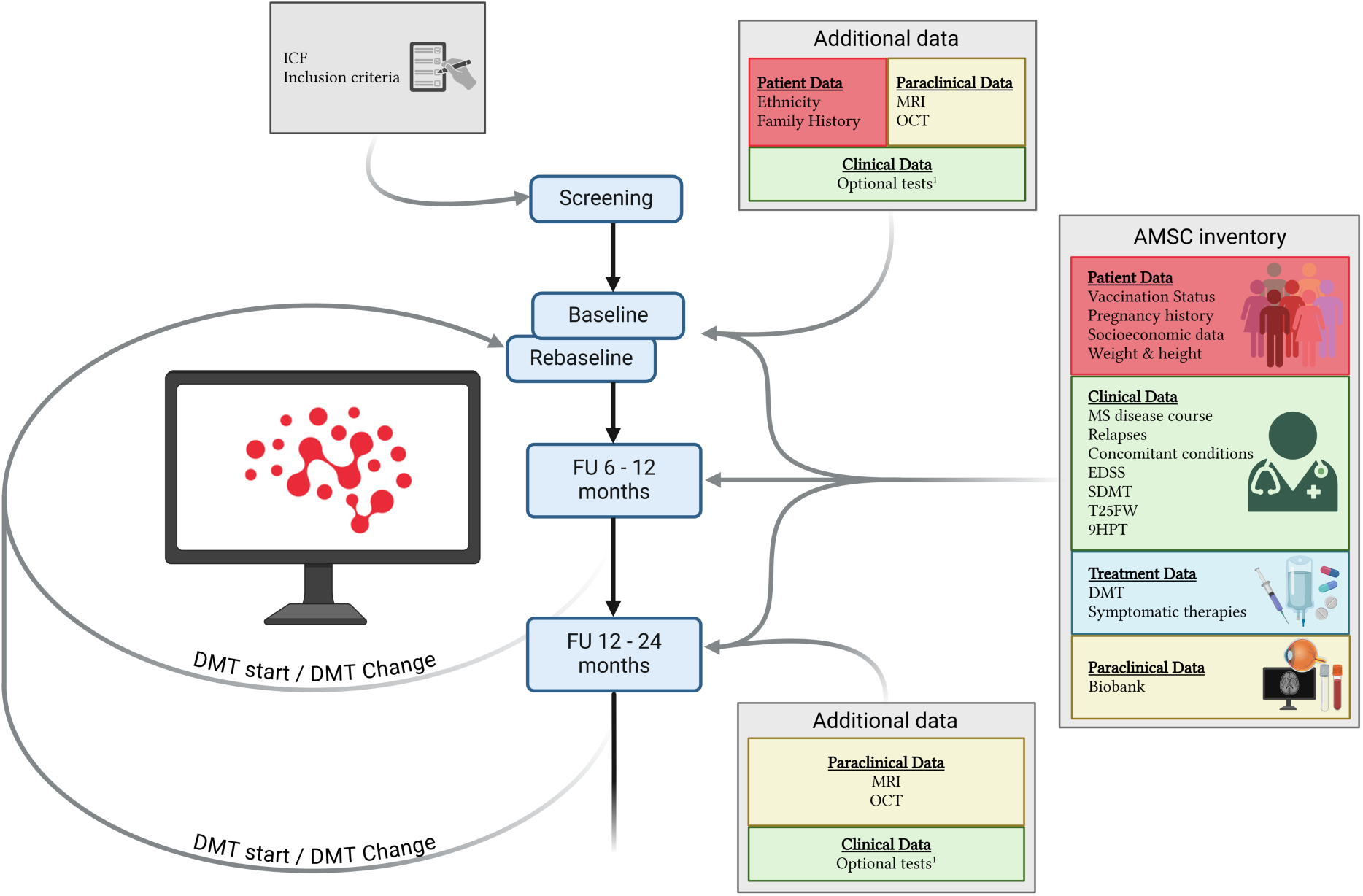
Overview Austrian MS Cohort. AMSC: Austrian Multiple Sclerosis Cohort. DMT: disease-modifying treatment. EDSS: expanded disability status scale. FU: Follow-up. ICF: informed consent form. MRI: magnetic resonance imaging. OCT: optical coherence tomography. SDMT: symbol digit modalities test. T25FW: Timed 25-foot walk test. 9HPT: 9-hole peg test. ^1^Optional tests include: Beck Depression Inventory (BDI), Fatigue Scale for Motor and Cognitive Fatigue (FSMC), Hospital Depression and Anxiety Scale (HADS), Multiple Sclerosis Impact Scale (MSIS-29), Personality, risk perception and coping in MS battery (PeRiCoMS), AMSC socioeconomic inventory (SCOPE-MS AT).

### Governance and Aggregated Data queries

The AMSD/AMSC Board, which consists of a fixed representative from each of the core centers in Vienna, Innsbruck, and Graz, along with two rotating representatives from participating centers (only one from each center allowed), oversees all aspects of conducting scientific nested research projects within the AMSD/AMSC.

Each project proposal must be submitted by the responsible project leader in writing to the AMSD/AMSC Board in the form of a brief protocol (including research question, inclusion and exclusion criteria, required data, and analysis plan) to. Each request is reviewed by the AMSD/AMSC Board. Projects require unanimous approval from the AMSD/AMSC Board.

The webRDA software enables cross-center aggregated data queries and analyses within the entire AMSD/AMSC. Centers not able to use the webRDA due to local legal or technical restriction can obtain data based on identical local queries. Each project proposal involving an aggregated AMSD/AMSC data query must be submitted to the Ethics Committee and the data-clearing committee of the Medical University of Vienna for review and approval. To ensure scientific quality as well as ethical and data protection integrity, aggregated database queries can only be conducted by the coordinating center at the Medical University of Vienna.

Upon approval by both the AMSD/AMSC Board and the Ethics Committee, the requested data are provided in pseudonymized form via secure data transfer (safe cloud or personal collection of a storage device) to the respective project leader. Project leaders are responsible for the ongoing security and integrity of the data, with any sharing or use outside the approved project strictly prohibited. Project leaders are also required to i) provide semi-annual progress reports and a final report on the analysis results, ii) submit any planned analyses based on AMSD/AMSC data, either partially or entirely, to the AMSD/AMSC board for review and approval prior to publication, iii) explicitly and visibly acknowledge the AMSD/AMSC as the data source in all publications derived partially or entirely from AMSD/AMSC data; and iv) recognize contributing MS centers in line with good scientific practice guidelines, either through acknowledgments or co-authorships.

### Perspectives

This project establishes the foundation for standardizing collection of comprehensive data of people with MS across Austria providing a data protection-compliant system for gathering population-based data.

Through a coordinated, consensus-driven approach, it substantially enhances the interconnectedness of MS centers throughout Austria.

Due to the inter- and intraindividual heterogeneity in MS course together with the ever-growing complexity of biomarker and treatment options, establishing an infrastructure for comprehensive harmonized collection of population-based data of people with MS is the most promising approach for facilitating evidence-based development of individualized treatment strategies. This project opens a myriad of scientific opportunities to analyze a plethora of diverse questions related to the diagnosis, treatment, and prognosis of people with MS, based on a high-quality data foundation.

For patients, benefits arise from both the high level of data security provided by the RDA system (including the webRDA application) and the improved quality assurance resulting from enhanced center connectivity as well as strengthened evidence-based practices, which will further advance the quality of clinical care.

Finally, by integrating into the RDA infrastructure – which is inherently scalable across the European Union – this project contributes to the creation of a digital ecosystem as part of a European Health Data Space for MS.

## Data Availability

All data produced in the present work are contained in the manuscript

## Funding

This project is funded by the Austrian MS Research Society.

## Acknowledgement

We thank all the AMSD/AMSC investigators, clinical research staff, and especially the patients for helping to collect these data. The named individuals are not compensated for their help.

## Group information AMSD/AMSC

### AMSD/AMSC investigators in alphabetical order

Altmann, Patrick (Medical University of Vienna, Vienna, Austria); Auer, Michael (Medical University of Innsbruck, Innsbruck, Austria); Berek, Klaus (Medical University of Innsbruck, Innsbruck, Austria); Berger, Thomas (Medical University of Vienna, Vienna, Austria); Bsteh, Gabriel (Medical University of Vienna, Vienna, Austria); Damulina, Anna (Medical University of Graz, Graz, Austria); Deisenhammer, Florian (Medical University of Innsbruck, Innsbruck, Austria); Demjaha, Rina (Medical University of Graz, Graz, Austria); Di Pauli, Franziska (Medical University of Innsbruck, Innsbruck, Austria); Enzinger, Christian (Medical University of Graz, Graz, Austria); Hegen, Harald (Medical University of Innsbruck, Innsbruck, Austria); Heschl, Bettina (Medical University of Graz, Graz, Austria); Hofer, Edith (Medical University of Graz, Graz, Austria); Kornek, Barbara (Medical University of Vienna, Vienna, Austria); Leutmezer, Fritz (Medical University of Vienna, Vienna, Austria); Martinez-Serrat, Maria (Medical University of Graz, Graz, Austria); Monschein, Tobias (Medical University of Vienna, Vienna, Austria); Pinter, Daniela (Medical University of Graz, Graz, Austria); Rommer, Paulus (Medical University of Vienna, Vienna, Austria); Ropele, Stefan (Medical University of Graz, Graz, Austria); Schmidauer, Martin (Medical University of Innsbruck, Innsbruck, Austria). Schmied, Christiane (Medical University of Vienna, Vienna, Austria); Tafrali, Cansu (Medical University of Graz, Graz, Austria); Wurth, Sebastian (Medical University of Graz, Graz, Austria); Zebenholzer, Karin (Medical University of Vienna, Vienna, Austria); Zinganell, Anne (Medical University of Innsbruck, Innsbruck, Austria); Zulehner, Gudrun (Medical University of Vienna, Vienna, Austria); Zrzavy, Tobias (Medical University of Vienna, Vienna, Austria).

## Disclosure of conflicts of interest

The AMSC is an investigator-initiated study with no financial interests.

**Gabriel Bsteh**: has participated in meetings sponsored by, received speaker honoraria or travel funding from Biogen, Celgene/BMS, Janssen, Lilly, Merck, Novartis, Roche, Sanofi-Genzyme and Teva, and received honoraria for consulting Biogen, Celgene/BMS, Janssen, Merck, Novartis, Roche, Sanofi-Genzyme and Teva. He has received unrestricted research grants from Celgene/BMS and Novartis. He serves as an Executive Committee of the European Committee for Treatment and Research in Multiple Sclerosis (ECTRIMS).

**Fabian Föttinger:** Has received speaker honoraria and travel funding from Novartis.

**Markus Ponleitner:** Has participated in meetings sponsored by, received speaker or consulting honoraria from Amicus and travel funding from Amicus, Merck, Novartis and Sanofi-Genzyme.

**Klaus Berek:** has participated in meetings sponsored by and received travel funding or speaker honoraria from Biogen, Merck, Novartis, Roche, Sanofi, and Teva. He is associate editor of Frontiers in Immunology / Neurology, Section Multiple Sclerosis and Neuroimmunology.

**Franziska Di Pauli:** has participated in meetings sponsored by, received honoraria (lectures, advisory boards, consultations) or travel funding from Bayer, Biogen, BMS/Celgene, Merck, Novartis, Sanofi, Roche, and Teva. Her institution has received research grants from Roche.

**Bettina Heschl:** has received funding for travel or speaker honoraria from Bayer, Biogen, Merck, Novartis, Roche, Sanofi-Genzyme and Teva.

**Sebastian Wurth:** has participated in meetings sponsored by, received honoraria or travel funding from Biogen, Merck, Novartis, Sanofi-Genzyme, Teva, Allergan, Ipsen Pharma and Roche.

**Florian Deisenhammer:** has participated in meetings sponsored by or received honoraria for acting as an advisor/speaker for Alexion, Almirall, Biogen, BMS, Sanofi, Amgen, Janssen, Laurea Group, Medwhizz, Merck, Novartis Pharma, Neuraxpharm, Roche, Sandoz, and Teva. His institution has received research grants from Biogen, Novartis Pharma, and Sanofi. He is section editor of the MSARD Journal (Multiple Sclerosis and Related Disorders) and review editor of Frontiers Neurology.

**Christian Enzinger:** has received funding for travel and speaker honoraria from Bayer, Biogen, Merck, Novartis, Roche, Sanofi-Genzyme, Shire and Teva. has received research support from Biogen, Celgene, Merck, and Teva; is serving on scientific advisory boards for Bayer, Biogen, Celgene, Merck, Novartis, Roche and Teva.

**Thomas Berger:** has participated in meetings sponsored by and received honoraria (lectures, advisory boards, consultations) from pharmaceutical companies marketing treatments for MS: Allergan, Bayer, Biogen, Bionorica, BMS/Celgene, Genesis, GSK, GW/Jazz Pharma, Horizon, Janssen, MedDay, Merck, Novartis, Octapharma, Roche, Sandoz, Sanofi-Genzyme, Teva and UCB. His institution has received financial support in the past 12 months by unrestricted research grants (Biogen, Bayer, BMS/Celgene, Merck, Novartis, Roche, Sanofi-Genzyme, Teva) and for participation in clinical trials in multiple sclerosis sponsored by Alexion, Bayer, Biogen, Merck, Novartis, Octapharma, Roche, Sanofi-Genzyme, Teva.

**Michael Khalil:** has received travel funding and speaker honoraria from Bayer, Biogen, Novartis, Merck, Sanofi and Teva and serves on scientific advisory boards for Biogen, BMS/Celgene, Gilead, Merck, Novartis, and Roche. He received research grants from Biogen, Novartis and Teva.

**Harald Hegen**: has participated in meetings sponsored by, received speaker honoraria or travel funding from Bayer, Biogen, Celgene, Janssen, Merck, Novartis, Sanofi-Genzyme, Siemens and Teva, and received honoraria for consulting Biogen, Celgene, Novartis, Roche, and Teva.

## Author contributions

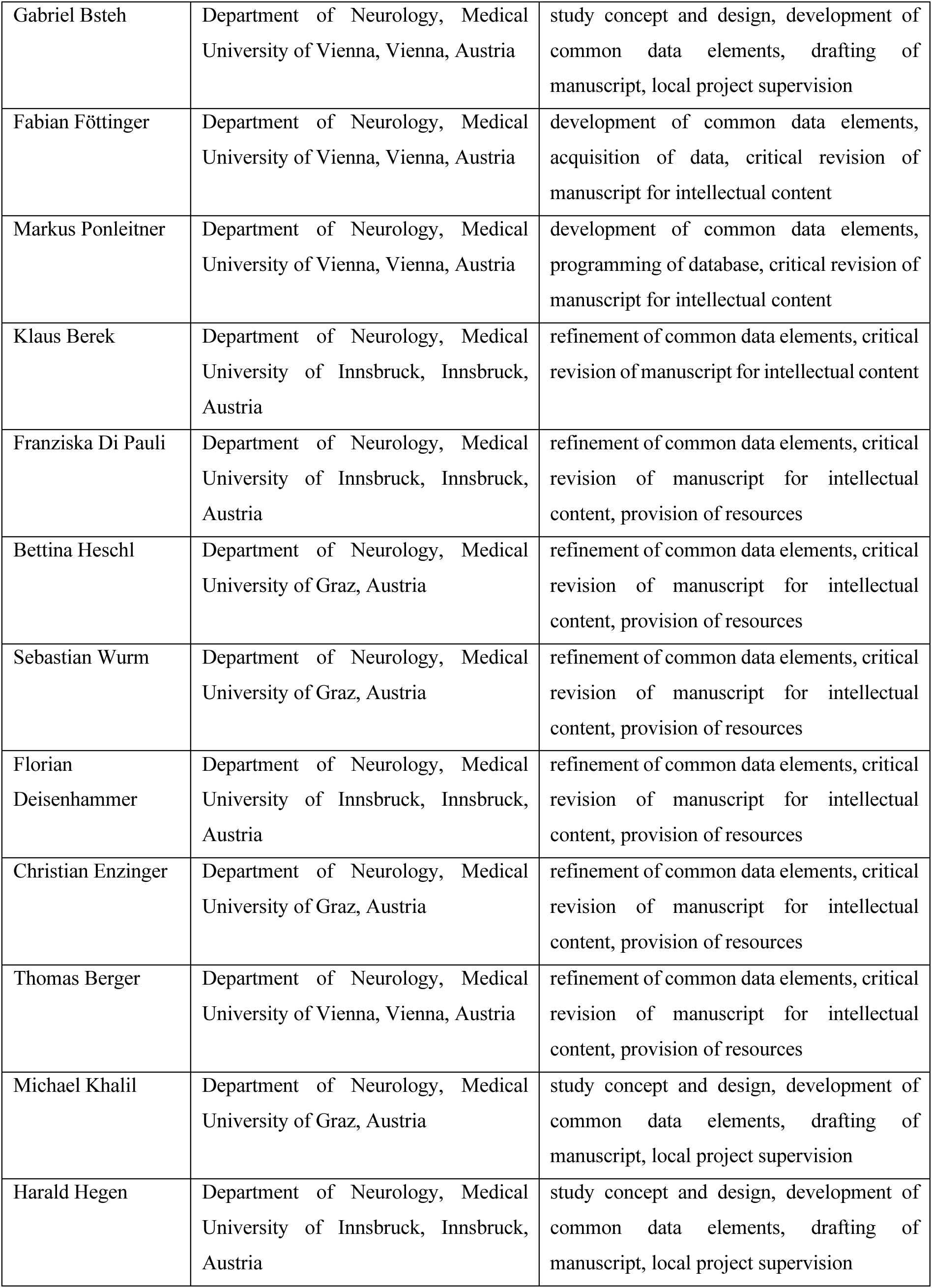

